# Distinct neural representations encode psychiatric states across timescales

**DOI:** 10.64898/2026.07.30.26359255

**Authors:** Timon Merk, Grace Bezold, Rahul Hingorani, Andrew D. Wiese, Brian Robinson, Han Yi, Jordan Altman, Sameer V. Rajesh, Victoria E. Gobo, Matthew Ochoa, Thomas Hamre, Vinayak Belavadi, Corey P. St. Romain, Michelle Avendano-Ortega, Sarah Soubra, Gabriel Reyes, Tomasz M. Frączek, Eric A. Storch, Wayne K. Goodman, Sameer A. Sheth, Nicole R. Provenza

## Abstract

Psychiatric states manifest over multiple timescales, from transient fluctuations in disorder-related distress to long-term variations in disease severity, yet whether these expressions share common neural representations remains unknown. We addressed this question in obsessive-compulsive disorder (OCD) by comparing neural activity associated with momentary distress evoked by triggers versus that associated with clinically assessed chronic symptom severity. We analyzed intracranial recordings from the ventral capsule and orbitofrontal cortex spanning more than 4595 patient-days in 8 individuals undergoing deep brain stimulation therapy (NCT03457675, NCT04281134, NCT04806516), together with quantitative facial- and speech-derived behavioral measures. Neural signatures of OCD severity were dissociable from and stronger than signatures of momentary distress, indicating distinct underlying brain states. Integrating behavioral data into a predictive computational model improved neural decoding performance of both momentary distress and OCD severity. These findings show that acute manifestations of psychiatric illness are not simply short-timescale readouts of the same neural states that encode longer-term clinical burden. Instead, transient distress and longitudinal disease severity are represented by dissociable neural processes. This distinction has important implications for how psychiatric biomarkers and adaptive neuromodulation strategies should be designed.

## Introduction

Psychiatric disorders are dynamic conditions in which clinically relevant states evolve across various temporal scales. Affected individuals may experience transient fluctuations in emotion, mood, or distress that occur within the context of slower changes in overall disease severity, functional impairment, and treatment response. A fundamental unresolved question is whether transient symptom expression reflects a rapidly varying readout of the same underlying neural state that determines longer-term disease burden, or whether these clinical dimensions are encoded by separable neural processes. Resolving this distinction is important both for understanding how psychiatric illness is organized in the brain and for determining which neural states should be targeted by adaptive neuromodulation or brain-computer interface therapies.

Obsessive–compulsive disorder (OCD) provides a particularly well-defined model in which to address this question. OCD is a chronic and debilitating psychiatric condition affecting approximately 2–3% of the global population^1,2^. Although individuals with OCD commonly experience episodes of acute distress elicited by exposure to environmental or internal triggers, the overall burden of the disorder is not clinically measured by these transient experiences but by the persistent inability to control obsessions and resist compulsions in everyday life. Consequently, successful treatment reflects more than reductions in momentary distress; it restores behavioral control, reduces avoidance, and diminishes interference of OCD symptoms with daily functioning. These latter dimensions are captured by the Yale-Brown Obsessive Compulsive Scale (Y-BOCS), the gold-standard measure of OCD symptom severity used to guide clinical decision-making and evaluate treatment efficacy. Accordingly, this scale quantifies these behaviors and effects over timescales of days to weeks^4,5^. Exposure and response prevention (ERP), the most effective behavioral treatment for OCD, exemplifies this distinction by requiring patients to repeatedly experience distress while learning to refrain from compulsive behaviors, thereby reducing long-term symptom severity without necessarily eliminating acute distress during individual exposures^6–8^. These clinical dissociations suggest that acute distress may represent a transient expression occurring within a broader disease state, rather than a direct moment-to-moment proxy for that state.

Prior neurophysiological studies of OCD have predominantly focused on neural activity measured during symptom provocation or task-based paradigms lasting seconds to minutes, with the goal of identifying neural biomarkers of acute distress, obsessive thoughts, or compulsive behaviors^9–12^. More recently, studies using on-device neural recordings have begun to identify neural correlates of longitudinal OCD symptom severity during DBS therapy over long timescales^13,14^, demonstrating that intracranial recordings can track disease progression over months to years of treatment. When considered in aggregate, however, this body of work has thus far failed to converge on a set of common neural biomarkers^15^. In contrast, the analogous pursuit in Parkinson’s disease has consistently highlighted subcortical and cortical neural features that track cardinal symptoms and can be used for closed-loop DBS control^16^.

One possible explanation for the heterogeneity in OCD is that acute distress and longitudinal disease severity represent distinct clinical constructs with different underlying neural representations. In support of this notion, clinical experience demonstrates that episodes of intense distress can occur in individuals with relatively mild OCD, whereas some individuals with severe OCD may experience periods of relatively little acute distress though remaining profoundly impaired by avoidance and diminished behavioral control. Recent advances in chronic DBS sensing provide a unique opportunity to determine whether neural representations of transient distress differ from those associated with longitudinal disease severity by measuring brain activity across the wide range of timescales over which these behaviors and symptoms evolve.

Although chronic neural recordings provide an opportunity to measure brain activity directly, psychiatric diagnosis and treatment remain fundamentally grounded in behavioral observation. Clinicians routinely integrate facial expression, speech, affect, and motor behavior when evaluating symptom severity and treatment response^17^. Advances in computational behavioral analyses now allow these observations to be quantified objectively from video and audio recordings^18–20^. Rather than serving solely as an additional source of clinical information, behavioral measurements may also guide the identification of neural features most relevant to disease state. By using these behavioral measures as supervisory signals, neural representation learning can incorporate the same types of clinically meaningful information that routinely inform psychiatric assessment. Using behavioral measurements as supervisory information might therefore enable neural representations that are more closely aligned with clinically relevant states^21^.

Here, we investigated whether momentary distress and longitudinal OCD symptom severity are represented by distinct neural signatures. Using chronic intracranial recordings acquired during DBS treatment, we separately characterized neural activity associated with naturalistic momentary distress during remote ERP teletherapy in patients’ homes and longitudinal fluctuations in clinical OCD symptom severity measured over hundreds of days. We further investigated whether objective facial and speech-derived behavioral measures could improve neural representations and decoding of these clinically relevant states. By distinguishing neural representations of transient distress from those associated with long-term disease severity, this work establishes a framework for developing neural decoders that are explicitly aligned with the clinical state they are intended to estimate. More practically, it provides a foundation for adaptive neuromodulation strategies that target the neural processes underlying long-term clinical improvement in psychiatric disorders.

## Results

### Chronic intracranial recordings during naturalistic exposures and longitudinal clinical follow-up

This study reports secondary, exploratory neurophysiological analyses of data acquired during a prospective early feasibility clinical trial to identify intracranial neural biomarkers of OCD (NCT03457675, NCT04281134 and NCT04806516). The three trial registrations correspond to successive device configurations: Activa PC+S (NCT03457675), Summit RC+S without electrocorticography (ECoG; NCT04281134), and Summit RC+S with orbitofrontal ECoG (NCT04806516). We implanted ten treatment-resistant OCD patients with DBS leads in the ventral capsule (VC) and connected the leads to investigational recording-capable DBS devices (Table 1). The present analyses included eight of these participants (P3– P10), who were implanted with the Summit RC+S recording-capable DBS system (participant flow diagram shown in Supplementary Figure 1). The first two participants (P1–P2) were implanted with the Activa PC+S system and were therefore excluded from these analyses. In a subset of patients (n=5), we additionally implanted bilateral orbitofrontal cortex (OFC) electrocorticography (ECoG) electrodes for passive neural sensing (P6-P10, Figure 1a). We previously reported two serious adverse events for the clinical trial^25^. P7 underwent mid-study IPG repositioning due to discomfort and migration and experienced a headache requiring admission, which self-resolved. In P10, a postoperative CT revealed a small intraparenchymal hemorrhage, which was asymptomatic and self-limited.

**Figure 1.**
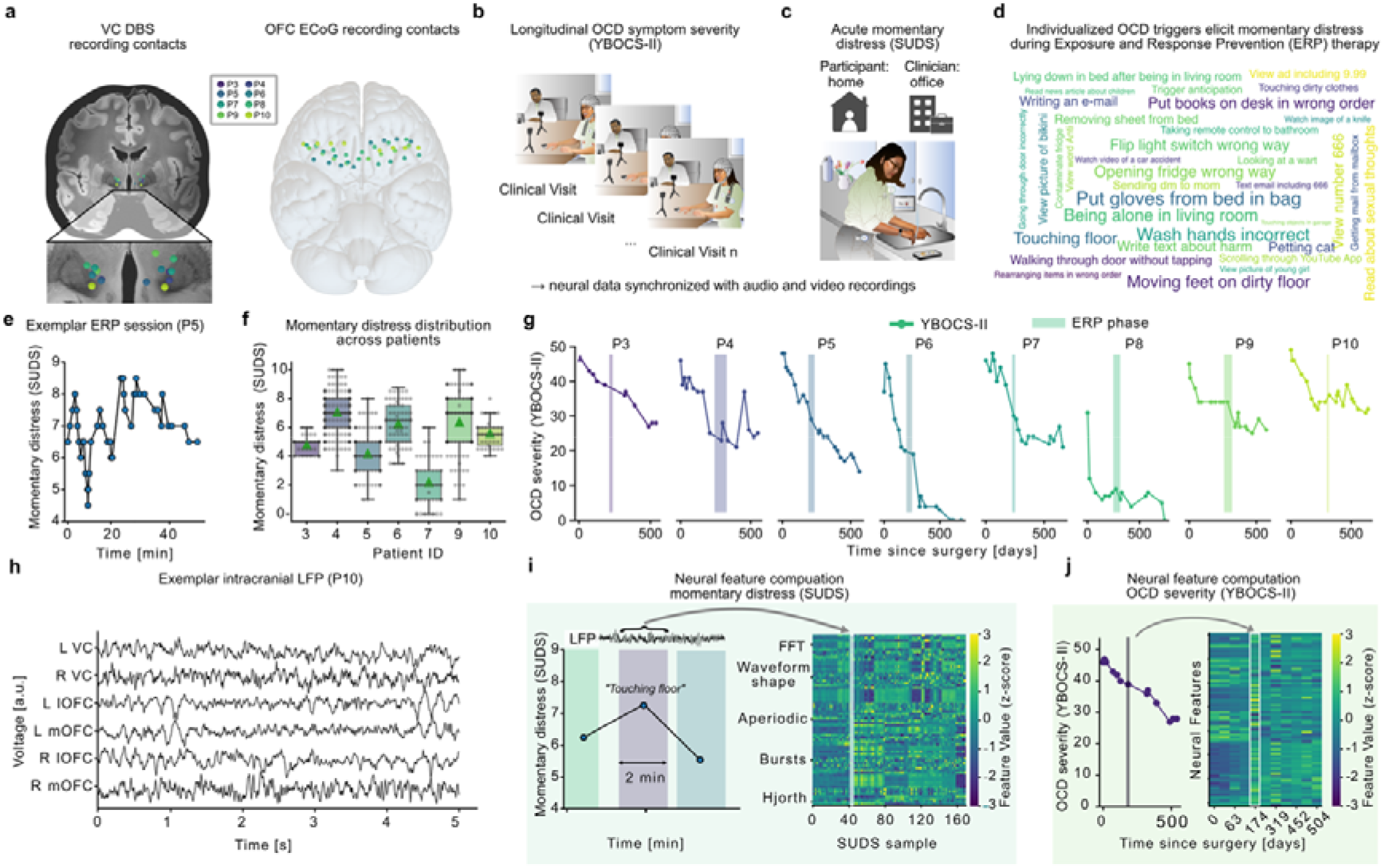
Experimental design for chronic intracranial recordings of longitudinal OCD symptom severity and momentary distress. **(a)** Bilateral ventral capsule (VC) DBS leads enabled chronic local field potential (LFP) recordings throughout an 18-month clinical trial. In five patients, bilateral orbitofrontal cortex (OFC) electrocorticography (ECoG) strip electrodes provided simultaneous cortical recordings. **(b)** OCD symptom severity was monitored longitudinally throughout the DBS clinical trial using the Yale-Brown Obsessive Compulsive Scale II (YBOCS-II). Figure adapted from^12^. **(c)** Exposure and response prevention (ERP) teletherapy sessions were conducted at patients’ homes via video conference. During ERP sessions, patients were exposed to individualized OCD-related triggers and reported momentary distress. Figure adapted from^12^. **(d)** ERP sessions enabled recording of neural activity during naturalistic, patient-specific OCD exposures encountered in everyday life. **(e)** Example ERP session illustrating within-session fluctuations in momentary distress. During ERP, participants were presented with symptom-provoking exposures and reported their Subjective Units of Distress Scale (SUDS) ratings, either when requested or when they experienced a change in distress. These assessments captured fluctuations in momentary distress over minutes within a single ERP session. **(f)** Across ERP sessions, repeated SUDS assessments yielded distributions of momentary distress ratings for each participant, representing short-timescale changes in affective state in contrast to **(g)** longer-timescale changes in OCD symptom severity over many months. **(h)** Neural features were extracted from time-series LFP recordings to identify neural correlates of **(i)** momentary distress and **(j)** longitudinal OCD symptom severity.

**Table 1:** Participant-level demographic and clinical characteristics of the eight participants included in the present secondary analysis. YBOCS = Yale-Brown Obsessive-Compulsive Scale; MDD = major depressive disorder; PTSD = post-traumatic stress disorder; GAD = generalized anxiety disorder; TS = Tourette Syndrome; IPH = intraparenchymal hemorrhage; M = male; F = female.

| Patient | Gender | Age (range) | Race | Ethnicity | Comorbid diagnoses | OCD subtypes | Age at OCD onset (range) | Initial YBOCS-II score | Final YBOCS-II score | Serious Adverse Events |
| --- | --- | --- | --- | --- | --- | --- | --- | --- | --- | --- |
| P3 | F | 36-40 | White | Hispanic/Latino | PTSD, MDD, TS | Harm, Not Just Right Experiences | 11-15 | 49 | 28 |  |
| P4 | M | 36-40 | White | Not Hispanic/Latino | MDD, Bipolar Disorder II | Checking, Contamination, Harm, Scrupulosity, Symmetry and Ordering | 16-20 | 42 | 25 |  |
| P5 | F | 31-35 | White | Not Hispanic/Latino | MDD | Contamination, Harm | 6-10 | 47 | 14 |  |
| P6 | M | 36-40 | Asian | Not Hispanic/Latino | GAD | Checking, Contamination | 26-30 | 37 | 0 |  |
| P7 | M | 36-40 | White | Not Hispanic/Latino | MDD | Harm, Not Just Right Experiences | 11-15 | 45 | 21 | Mid-study IPG repositioning due to discomfort and migration. Headache requiring admission; self-resolved. |
| P8 | F | 36-40 | White | Not Hispanic/Latino | MDD, GAD | Checking, Contamination, Harm, Not Just Right Experiences | 6-10 | 37 | 1 |  |
| P9 | F | 26-30 | White | Hispanic/Latino | MDD, GAD | Contamination, Harm, Not Just Right Experiences, Scrupulosity | 11-15 | 45 | 26 |  |
| P10 | M | 31-35 | White | Not Hispanic/Latino | MDD | Checking, Contamination, Not Just Right Experiences | 11-15 | 46 | 32 | Postoperative CT revealed small IPH at the inferior left frontal lobe, patient remained asymptomatic and the IPH was self-limited |

Throughout the 18-month clinical trial, patients underwent repeated assessments of OCD symptom severity during clinic visits (22.5±1.5 assessments per patient; Figure 1b; complete trajectory of YBOCS-II scores including assessments without corresponding neural recordings is shown in Supplementary Figure 2). Patients additionally participated in ERP teletherapy sessions in their homes (6.6±3.4 sessions per patient; P6 was excluded from ERP analysis due to neural artifact contamination, Figure 1c). During ERP, patients were exposed to personalized OCD triggers (Figure 1d) and verbally reported momentary distress from zero to ten using the subjective units of distress rating scale (SUDS; ratings per session: 12.3±8.5; Figure 1e-f). All but one patient achieved a clinically significant response to DBS (responder criteria: ≥35% reduction in YBOCS^26^; Table 1), resulting in substantial longitudinal variation in OCD symptom severity throughout the study^25^ (Figure 1g).

These complementary paradigms enabled us to examine neural activity associated with two clinically relevant but distinct phenomena: momentary distress experienced during individualized ERP exposures and fluctuations in longitudinal OCD symptom severity (Figure 1h-j). Therefore, we recorded local field potentials (LFP) bilaterally from the VC DBS target (n=8) and the OFC (n=5) during both clinic visits and at-home ERP sessions (Figure 1h). From these recordings, we extracted a comprehensive set of spectral, temporal, waveform, bursting, and connectivity features (Methods; Figure 1i,j)^24,27^. To investigate neural features associated with momentary distress, we analyzed the neural recording surrounding each SUDS rating collected during ERP, and for long-term OCD symptom severity, we computed one neural feature vector per clinical visit and paired it with the corresponding YBOCS-II score obtained at that visit. This experimental design allowed us to directly compare neural representations of transient distress and longitudinal clinical symptom severity within the same individuals.

### Distinct neural representations of momentary distress and long-term symptom severity

We first asked whether longitudinal OCD symptom severity and momentary distress exhibit similar neural correlates. To address this question, we computed correlations between a comprehensive set of intracranial neural features and both OCD symptom severity and momentary distress across all recorded brain regions (Figure 2a-b, Supplementary Figure 3). Motivated by previous findings implicating cortico-basal hyperconnectivity in OCD^10,28,29^, we tested whether OFC–VC connectivity was associated with OCD symptom severity. In two of five patients, delta coherence across bilateral OFC–VC channels was significantly and positively correlated with symptom severity (Supplementary Figure 3). At the group level, this pre-specified hypothesis test also showed a significant positive association (*R*=0.14±0.29, *p*=0.039), although it did not remain significant after correction for multiple comparisons (corrected *p*=0.13, Supplementary Figure 3).

**Figure 2.**
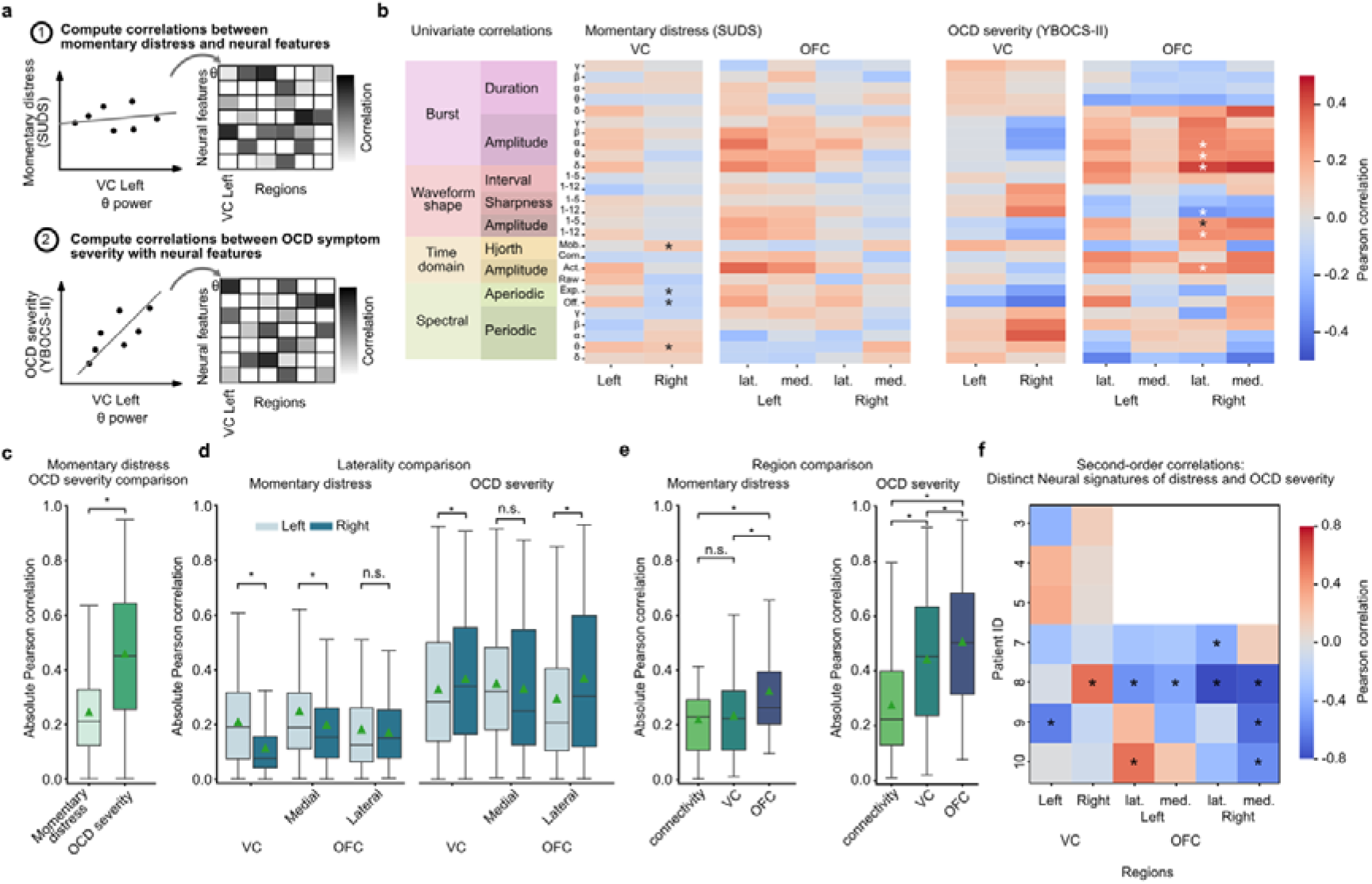
Neural signatures of momentary distress and longitudinal OCD symptom severity. **(a)** Overview of correlation analyses comparing neural features with momentary distress and OCD symptom severity. **(b)** Mean region- and hemisphere-specific Pearson correlation heatmaps across patients, including orbitofrontal cortex (OFC) and ventral capsule (VC) recordings in both left and right hemispheres. Significant correlations after multiple-comparison correction are marked with (*). Patient-specific correlations are shown in Supplementary Figure 3. **(c)** Absolute neural feature correlations with OCD symptom severity and momentary distress. **(d)** Comparison of left and right hemisphere correlations for momentary distress and OCD symptom severity separate for VC, lateral and medial OFC. **(e)** Comparison of VC, OFC and connectivity correlation strengths for momentary distress and OCD symptom severity. **(f)** Second-order correlation analysis comparing feature–region correlation maps for momentary distress and OCD symptom severity for VC and OFC recordings.

Across all features, neural activity demonstrated significantly stronger associations with longitudinal OCD symptom severity than with momentary distress (|*R|=*0.46±0.23 vs. |*R|=*0.24±0.17, respectively; relative permutation test *p=*2·10^−4^, mean participant-level Δ|*R*|=0.19±0.15, 95% CI [0.07, 0.30], participant-level permutation test *p*=0.02, 6/7 participants had stronger neural correlations with OCD symptom severity, Supplementary Table 2, Figure 2c). Furthermore, neural correlates of momentary distress exhibited significant lateralization within the VC and medial OFC, with stronger associations observed in the left hemisphere than the right (relative permutation test VC *p*=0.003; medial OFC *p*=0.001, lateral OFC *p*=0.51, Figure 2d). In contrast, neural correlates of OCD symptom severity showed significantly stronger associations in the right than the left VC and lateral OFC (relative permutation test VC *p*=0.027; lateral OFC *p*=0.0006, Figure 2d). OFC demonstrated significantly stronger correlations than VC for both OCD symptom severity (OFC |*R|=*0.50±0.24, VC |*R|=*0.36±0.21, relative permutation test *p*=0.0002, mean participant-level difference Δ|*R*|=0.14±0.08, 95% CI [0.07, 0.21], participant-level permutation test *p*=0.05, neural correlations were stronger in OFC for 5/5 patients) and momentary distress (OFC |*R|=*0.32±0.18, VC |*R|=*0.22±0.16, relative permutation test *p*=2·10^−4^, mean participant-level difference Δ|*R*|=0.10±0.09, 95% CI [0.02, 0.20], participant-level permutation test *p*=0.06, neural correlations were stronger in OFC for 4/4 patients, Supplementary Table 3, Figure 2e). Together, these findings show that the neural representation of symptom severity is more strongly represented than that of momentary distress; that these neural representations are lateralized; and that cortical features correlate with both severity and distress more strongly than do subcortical features.

Because DBS parameters were adjusted throughout the clinical trial, we next asked whether the observed neural correlates of OCD symptom severity could simply reflect acute stimulation effects rather than longitudinal changes in clinical state (Supplementary Figure 4). To control for this effect, we used neural recordings acquired before and after stimulation parameter changes within individual visits to distinguish acute programming-related changes in neural features from longitudinal changes associated with fluctuations in OCD symptoms. Although DBS programming produced measurable changes in neural features (Supplementary Figure 5), these changes were significantly smaller than those observed longitudinally across visits (Bernoulli test comparing the number of feature changes across versus within visits: *p*=1.5·10^−17^). These findings support that the observed neural correlates primarily reflected longitudinal changes in OCD symptom severity rather than the amount of stimulation delivered or acute effects of DBS programming.

Finally, we asked whether the neural representations of momentary distress and longitudinal OCD symptom severity captured a common underlying neural organization. To address this question, we computed second-order correlations that quantified the similarity between the feature-region correlation maps of OCD severity and momentary distress. In the VC, second-order correlations were centered near zero (*R*=−0.02±0.29, 95% CI [-0.17, 0.13]), with moderate evidence favoring the null hypothesis of no systematic correspondence (BF_01_=3.62, permutation *p*=0.82, Figure 2f). In the OFC, second-order correlations showed a significant negative relationship (*R*=−0.28±0.37, 95% CI [−0.46, −0.09], permutation test *p*=0.008). Together, these findings show that the neural representations of momentary distress and longitudinal OCD symptom severity are independent (in the VC) or even negatively correlated (in the OFC).

### Quantification of facial and speech features across momentary and longitudinal clinical states

Having established that momentary distress and longitudinal OCD symptom severity are associated with distinct cortical and subcortical neural signatures, we therefore quantified objective facial and speech-derived behavioral features across both clinical states. We extracted facial action units (FAUs)^32^ and patient-derived acoustic speech features (OpenSmile acoustic metrics^30^, Wav2Vec2 speech embeddings^33^, and valence, arousal, dominance speech estimates^31^) from video and audio recordings obtained during clinic visits and ERP sessions (Figure 3a). For momentary distress, video- and audio-derived behavioral features were averaged surrounding each SUDS rating during ERP sessions. Likewise, for longitudinal OCD symptom severity, behavioral features were averaged across each clinical interview and paired with the corresponding YBOCS-II score. In this way, we objectively quantified patterns of facial expressions and verbal output produced by each patient across various states of momentary distress and of clinical response. Behavioral correlates of both measures were highly individualized across participants (Figure 3b), with participant-specific structure visible in the low-dimensional FAU and speech representations (Figure 3c-d).

**Figure 3.**
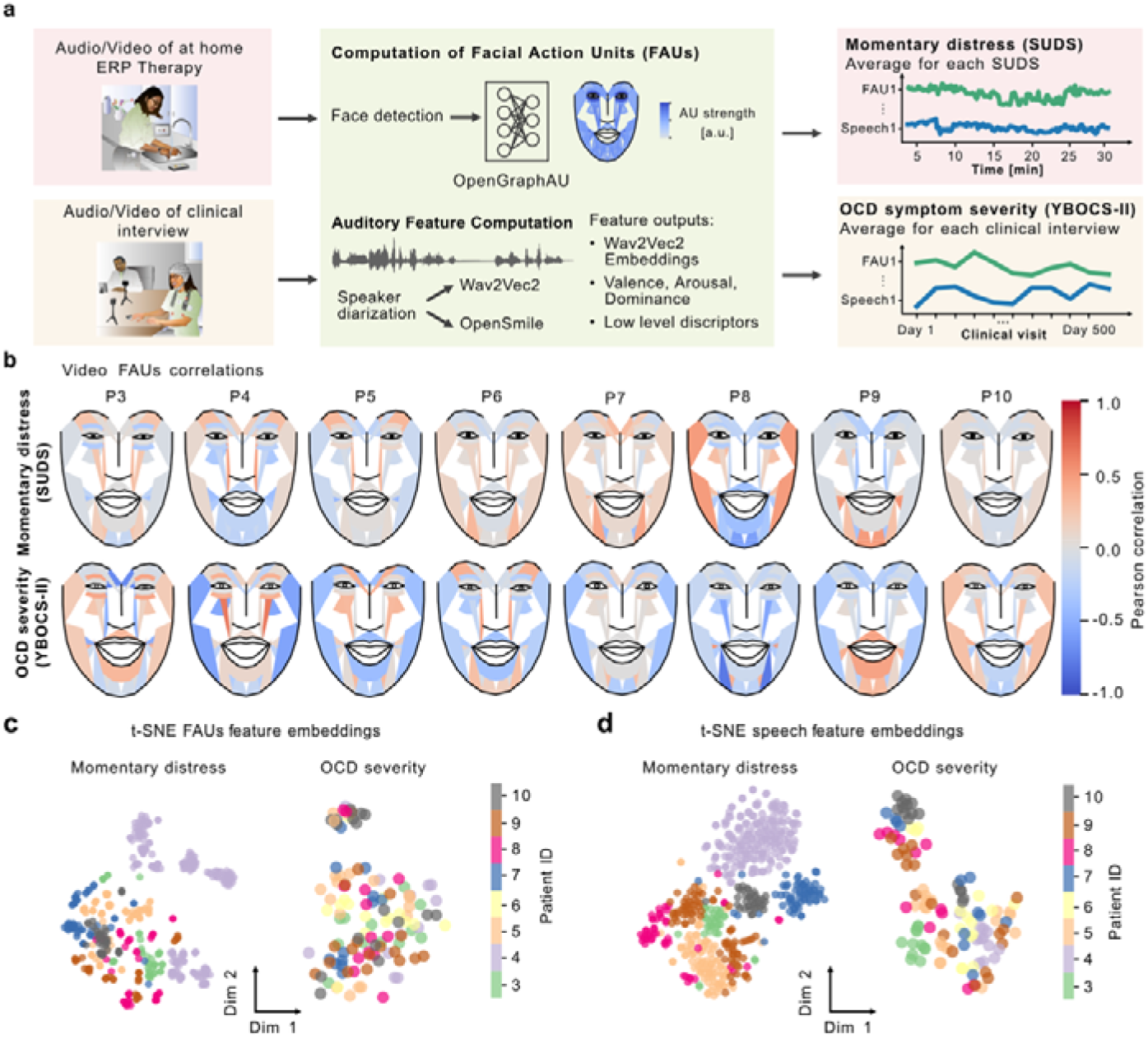
Quantification of facial and speech features across momentary and longitudinal clinical states. **(a)** Multimodal analysis integrating chronic intracranial neural recordings with facial expression and speech characteristics extracted from video recordings obtained during at-home exposure and response prevention (ERP) therapy and interviews during clinic visits. Facial action units (FAUs) and speech characteristics (OpenSmile derived acoustic features^30^, Wav2Vec2 embeddings and valence/arousal/dominance estimates^15,31^) were extracted from video and audio recordings, respectively. For momentary distress, behavioral features were averaged over windows around each distress rating. For OCD symptom severity, behavioral features were averaged within each clinical interview and paired with the neural recording corresponding to each YBOCS-II assessment. Figure adapted from^12^. **(b)** Patient-specific correlations between FAUs and momentary distress (top) or OCD symptom severity (bottom). **(c)** Patient-specific t-SNE visualization of FAUs and **(d)** speech features with momentary distress and OCD symptom severity.

### Joint neural–behavioral modeling enhances OCD symptom decoding

Given the significant but distinct neural and behavioral correlates of acute vs. chronic measures, we next tested whether these correlates were clinically relevant and complementary. We assessed clinical relevance by incorporating these variables within a predictive modeling framework to determine their ability to predict, rather than simply correlate with, measured outcome. To assess complementarity, we compared the performance of models utilizing subsets of the neurobehavioral data as predictors. We hypothesized that neural and behavioral data provide independent information regarding clinical state (i.e., acute distress and chronic severity), such that models trained using the full neurobehavioral dataset would outperform models trained on a single modality.

To do so, we leveraged the CEBRA framework^34^ to learn latent neural representations using either neural features alone or neural features jointly trained with behavioral auxiliary variables (Figure 4a). Importantly, behavioral variables were used only during latent representation learning to shape the latent neural embedding; all decoding performance was subsequently evaluated using neural recordings alone, such that test set predictions require only neural data and do not depend on continuous monitoring of behavioral markers. In the neural-only model, latent embeddings were learned using the temporal structure of neural activity alone, whereas the behavior-guided model used facial and speech features to organize neural samples with similar behavioral representations. We hypothesized that, if the behavioral data provided information about the outcome measures beyond that captured by the neural data, incorporating these data would increase the separability of the learned representations and thereby improve outcome prediction. We trained separate models to decode momentary distress and longitudinal OCD symptom severity within each participant using leave-one-sample-out cross-validation. For decoding OCD symptom severity, we left out the entire recording from each clinical visit for test evaluation. This framework enabled us to directly compare neural-only embeddings with joint neural-behavioral embeddings and determine whether behavioral information improves downstream neural decoding.

**Figure 4.**
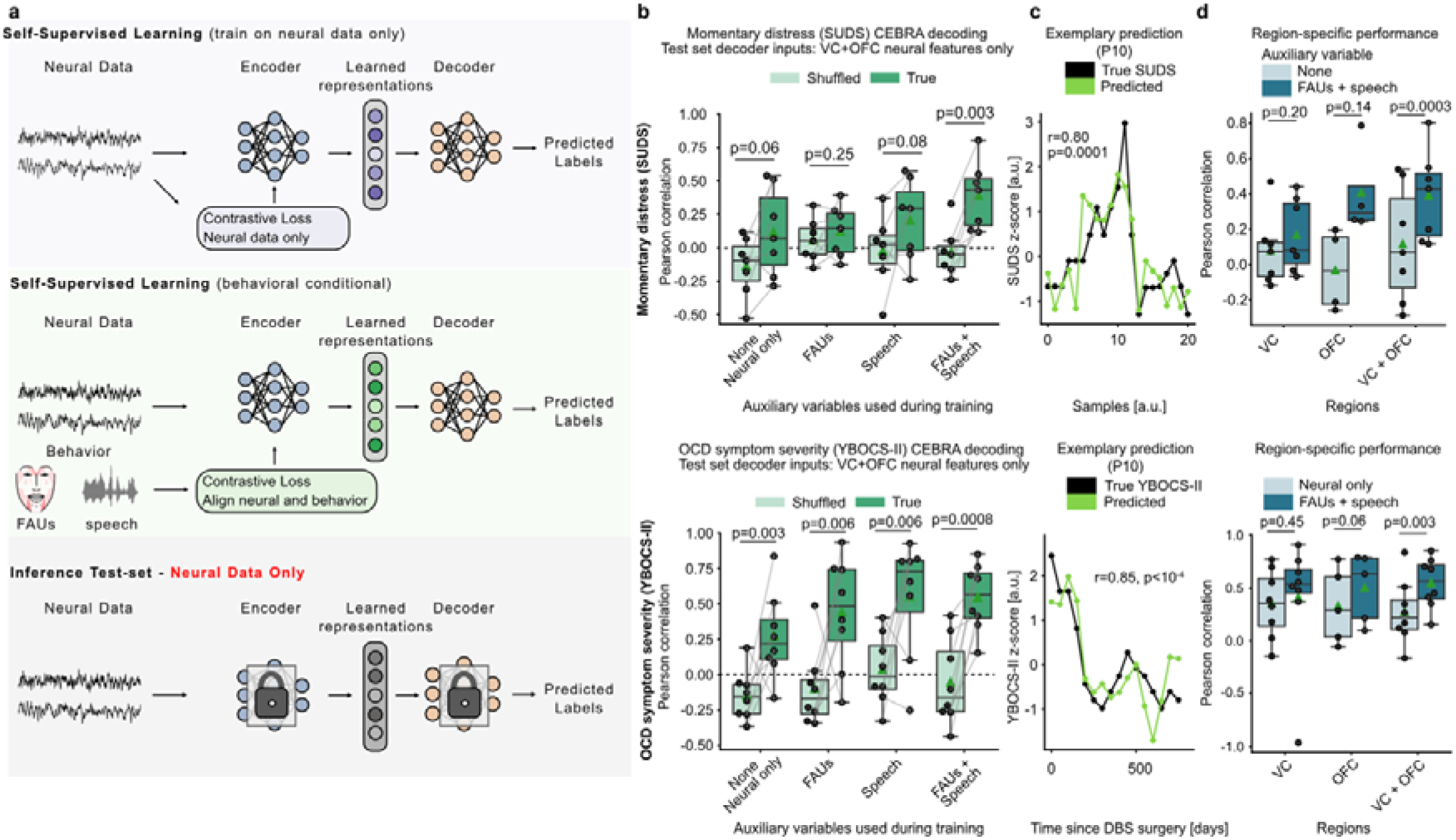
Behavior-guided neural representation learning improves decoding of momentary distress and longitudinal OCD symptom severity. **(a)** Overview of the CEBRA-based^34^ representation learning and decoding framework. Neural embeddings were learned either from neural recordings alone (self-supervised) or jointly with auxiliary behavioral variables (facial action units and speech features). During self-supervised training, neural samples were grouped solely based on temporal proximity (top). In contrast, during behavior-guided training, neural samples with similar behavioral representations were grouped within the latent space (middle). During inference, only neural recordings were provided to the frozen encoder and downstream decoder (bottom). **(b)** Decoding performance for momentary distress (top) and OCD symptom severity (bottom) using neural-only and behavior-guided neural embeddings. Chance-level decoding performance was estimated using shuffled clinical labels. **(c)** Representative predictions of momentary distress (P10; top) and OCD symptom severity (P10; bottom) using neural-only and behavior-guided embeddings. **(d)** Regional decoding performance using ventral capsule (VC)-only, orbitofrontal cortex (OFC)-only, or combined VC-OFC neural features.

Joint neural-behavioral representation learning did indeed enable improved decoding performance compared to neural-only models for both measures (Figure 4b-c). For acute distress (i.e., predicting SUDS ratings), neural-only latent embeddings performed at chance levels (actual *R*=0.12±0.31, shuffled *R*=-0.14±0.02, *p*=0.06), but the model achieved significant decoding performance with inclusion of behavioral variables (actual *R*=0.39±0.23, shuffled *R*=-0.03±0.17, *p*=0.003). Across the board, decoding of chronic symptom severity outperformed decoding of momentary distress, consistent with the stronger neural correlations for the former (Figure 2c). For chronic severity (i.e., predicting YBOCS-II scores), neural-only latent embeddings performed above chance (actual *R*=0.27±0.28, shuffled *R*=-0.15±0.16, *p*=0.003), but performance improved significantly when facial and speech variables were included (actual *R*=0.55±0.22, shuffled *R*=-0.06±0.28, *p*=0.0008). Notably, behavior-guided representation learning significantly improved decoding for momentary distress by Δ*R*=0.27±0.24 (participant-level bootstrap 95% CI [0.13, 0.45], improvement observed in 7/7 participants) and for OCD symptom severity by Δ*R*=0.27±0.27 (participant-level bootstrap 95% CI [0.11, 0.45], improvement observed in 7/8 participants, Supplementary Table 4). These findings demonstrate that face- and speech-derived behavioral features provide independent and complementary predictive information regarding clinical states beyond that provided by cortical and subcortical intracranial neural recordings. Moreover, this contrastive learning approach allows incorporation of the rich behavioral data into the learned low-dimensional representation when they are available during model training but does not require them during inference. Since the neural data are passively recorded on the DBS device and therefore readily available, but the behavioral data require availability of external audio/video equipment, this approach provides enhanced decoding performance even when only neural data are available during the vast majority of the time in real-world settings.

We further investigated the relative model contributions of neural data from the cortex (OFC) vs. the subcortical stimulation site (VC). Based on the correlation analyses above (Figure 2e) and results from other conditions such as Parkinson’s disease^35–37^, we hypothesized that cortical signals would enhance decoding. We therefore assessed decoding performance by restricting neural features to VC-only, OFC-only, or combined VC-OFC features. OFC-only embeddings yielded the highest decoding performance for acute distress (VC: *R=*0.17±0.21, OFC: *R=*0.41±0.26, VC-OFC: *R=*0.39±0.25), whereas combining VC and OFC produced the highest decoding performance for chronic symptom severity (VC: *R=*0.41±0.58, OFC: *R=*0.50±0.33, VC-OFC: *R=*0.55±0.24; Figure 4d).

Finally, we asked whether decoders optimized for one clinical state generalized to the other. Consistent with the second-order correlation results (Figure 2f), decoders trained to predict acute distress failed to predict chronic symptom severity, and vice versa. Specifically, distress-trained decoders showed significantly reduced performance when applied to chronic symptom severity compared with severity-trained decoders (mean patient-level Δ*R*=0.47±0.49, 95% CI [0.25, 0.85], 7/7 patients, permutation test *p*=0.01). Likewise, severity-trained decoders showed significantly reduced performance when applied to distress compared with distress-trained decoders (mean patient-level Δ*R*=0.71±0.42, 95% CI [0.43, 0.98], permutation test *p*=0.01; 7/7 patients, Supplementary Figure 6). These cross-decoding results provide convergent evidence that acute distress and chronic symptom severity are encoded by distinct neural representations.

## Discussion

Our findings reveal that an acute manifestation of psychiatric illness is not simply a short-timescale expression of the same neural state that encodes longer-term clinical burden. In OCD, momentary distress and longitudinal symptom severity were associated with dissociable neural representations, and neural decoders trained on one state failed to generalize to the other. These results suggest that psychiatric illness may be organized across nested but separable neural states, with transient symptom expression occurring within broader disease-related dynamics.

This distinction has important implications for the interpretation of psychiatric biomarkers. Momentary distress is inherently transient, context-dependent, and typically assessed through subjective self-report, whereas symptom severity is measured using clinical scales acquired by trained assessors that capture a broader disease state incorporating chronic burden. Notably, this dissociation existed even though the distressing exposures occurred naturalistically in the home environment, indicating that acute distress and clinical severity remain separable even under ethologically relevant symptom-provoking conditions. Because the YBOCS-II assesses the ability to tolerate obsessions, resist compulsions, limit avoidant behaviors, and maintain behavioral control over a timescale of weeks, our findings suggest that longitudinal clinical measures capture aspects of disease state not reflected by isolated distress ratings. More broadly, these findings suggest that clinically relevant psychiatric states are organized across multiple temporal scales, with distinct neural representations encoding different aspects of disease. Accordingly, psychiatric biomarkers should be developed with careful consideration of the clinical state they are intended to estimate.

Our findings further demonstrate that objective behavioral measures provide complementary information that enhances neural representations of clinically relevant psychiatric states. Clinical assessment in psychiatry relies heavily on behavioral observation, including facial expression, speech, affect, and motor behavior, to infer symptom severity and treatment response^17^. Consistent with this clinical practice, previous studies have demonstrated the value of behavioral readouts for quantifying psychiatric disorders^18–20,38,39^, while separate work has shown that behavior-guided latent representation learning can improve downstream decoding^34^. Our findings extend these observations by demonstrating that objective behavioral measures can improve neural representations of both short-term distress and long-term symptom severity, suggesting that behavioral information can guide the identification of clinically meaningful neural states.

Importantly, behavioral information was used only during latent representation learning and was not required during model inference. Rather than functioning as additional decoder inputs, behavioral measures improved the organization of the latent neural representation itself, resulting in substantially improved decoding of both clinical measures. One explanation for this improvement is that intracranial neural recordings simultaneously reflect numerous ongoing brain processes, many of which are unrelated to the clinical state of interest. By providing an independent measure of clinically relevant behavior, facial and speech features may guide representation learning toward neural dimensions that preferentially encode disorder-related neurophysiological processes while reducing emphasis on unrelated neural variability.

The improved decoding performance despite modest bivariate correlations between behavioral features and OCD symptom severity (Figure 3d) may reflect nonlinear relationships captured by CEBRA’s latent space. By conditioning the latent embedding on behavioral features during behavioral contrastive learning, CEBRA can reveal neural patterns that become informative for symptom decoding only in specific behavioral contexts or through nonlinear interactions^34^, even when either modality shows little linear association with symptom severity in isolation.

Our findings suggest that objective behavioral measures can provide meaningful supervisory information for learning neural representations of psychiatric state while preserving the ability to decode from neural activity alone. This arrangement is critical, as behavioral data are often the bottleneck, whether acquired from external cameras and microphones (which are intrusive and limited in their deployability) or from clinical interactions (which are relatively infrequent due to patient- and provider-related practical limitations). As methods for joint neuro-behavioral pre-training continue to advance^21^, this framework will enable predictive models to be pre-trained on large multimodal datasets and subsequently fine-tuned using sparser psychiatric ratings, thereby facilitating robust biomarker discovery and longitudinal prediction of disease state without increasing cost and burden to practitioners and patients.

Our findings also provide insight into the neural circuitry underlying OCD symptom expression and its implications for closed-loop or adaptive DBS. According to the prevailing model of OCD pathophysiology, symptoms arise from dysfunction within cortico-striato-thalamo-cortical circuits that regulate reward-motivated behavior and controlled decision-making^10,13,40^. Consistent with this framework, we found that elevated delta-band coherence between cortical and striatal regions was associated with greater OCD symptom severity (Supplementary Figure 3), suggesting that worsening clinical state is accompanied by alterations in circuit-level communication. Interestingly, univariate cortical signals consistently exhibited stronger associations with longitudinal OCD symptom severity than either subcortical or connectivity measures, consistent with the hypothesis that cortical recordings provide particularly rich information about disease state. Aligned with this hypothesis, cortical signals have shown stronger predictive power for decoding movement and symptom severity in Parkinson’s disease^35,36^. These findings suggest that future adaptive DBS systems may benefit from incorporating cortical recordings as biomarker sources^41^. More generally, these findings illustrate the need to separate priorities of optimizing therapeutic stimulation sites (which are often subcortical to engage broadly distributed networks) from neural biomarker recording sites (which may benefit from the specificity of cortical signals) to facilitate the development of more effective closed-loop neuromodulation systems.

The delayed therapeutic effects of DBS for psychiatric disorders provide important insight into the mechanisms underlying therapeutic response in psychiatric neuromodulation. Unlike in Parkinson’s disease, where stimulation often produces rapid symptomatic benefit, clinical improvement after DBS for OCD typically emerges only after weeks to months of DBS therapy^42^. Rather than acutely suppressing obsessions, VC stimulation may facilitate a gradual learning process that helps patients progressively tolerate and manage OCD-related distress over time^7,8,13^. This proposed mechanism parallels ERP, in which repeated exposure to symptom-provoking triggers while refraining from engaging in compulsions gradually strengthens distress tolerance and behavioral control rather than immediately eliminating distress. Our clinical experience similarly suggests that obsessive thoughts rarely disappear during the initial months of DBS treatment. Instead, patients become increasingly able to tolerate obsessive thoughts and resist compulsive urges^25,43^. This gradual trajectory of recovery is consistent with our correlation and prediction findings, which show that longitudinal clinical severity is more robustly represented by neural activity than is transient distress. These longitudinal neural signatures may therefore provide a window into the slowly evolving neural state that changes at a rate similar to therapeutic response after DBS. This observation raises the possibility that psychiatric DBS may exert its therapeutic effects by gradually modifying an underlying disease-related state rather than by acutely suppressing individual episodes of distress.

The concept that neuromodulation exerts its effect by decreasing the likelihood of pathological manifestations of a disorder rather than disrupting individual bouts of symptom expression is exemplified in epilepsy. Episodic seizure events occur within the background of slower-varying neural states that influence seizure susceptibility^44–46^. Notably, open-loop DBS can reduce seizure burden without necessarily interrupting individual seizures as they occur. Recent evidence suggests that even responsive neurostimulation, which is conceptually designed to abort individual seizure events, may exert its therapeutic effect by influencing seizure likelihood over longer timescales. More broadly, these observations support a framework in which transient pathological events do not necessarily constitute direct readouts of the slower neural states that determine susceptibility to those events.

Although episodes of momentary distress in OCD are fundamentally different from epileptic seizures, our results suggest that a similar distinction between background disease state and transient symptom expression may exist. We hypothesize that longitudinal OCD symptom severity reflects a slowly varying neural state that shapes how patients respond to everyday triggers, whereas momentary distress represents a transient behavioral manifestation occurring within that broader disease state^38,47–49^. Under this framework, the brain states captured by chronic neural recordings, which reflect patients’ ability to tolerate distress, resist compulsions, and adapt habitual behaviors^7,8^ are more saliently represented in cortical and subcortical neural activity than those related to transient episodes of momentary distress. Future studies incorporating continuous neural sensing and longitudinal behavioral monitoring are needed to further test this hypothesis.

Our dataset includes a large and robust collection of intracranial cortical and subcortical time-domain neural recordings acquired in the home environments of OCD patients during naturalistic exposures and across clinical visits, spanning more than 4595 patient-days. Despite this extensive dataset, both the number of participants and the number of independent distress and symptom ratings remain limited relative to the neural data. Given the substantial heterogeneity in symptom expression and neural biomarkers observed across psychiatric disorders^15^, we therefore developed within-patient decoders. As larger datasets become available, more complex models may become feasible and enable generalization across patients^24,35^. Similar to neuroimaging research, establishing robust electrophysiological signatures will require multi-cohort validation and large-scale dataset harmonization^50^.

An important future implication of this framework is using it to determine whether adaptive neuromodulation should target slowly evolving disease states rather than transient symptom events. If psychiatric disorders are organized across varying neural timescales, future closed-loop therapies may benefit from modulating the underlying clinical state that governs long-term clinical recovery instead of reacting to individual symptom episodes. This control objective differs from that of adaptive DBS in Parkinson’s disease, where stimulation can be adjusted in response to neural signals that track motor symptoms over relatively short timescales. Because therapeutic effects in psychiatric DBS emerge more gradually, adaptive systems may instead need to monitor and stabilize slower-varying clinical states while maintaining stimulation sufficient to sustain therapeutic circuit dynamics. If confirmed, it could fundamentally shift the design of adaptive neuromodulation from reactive symptom detection toward modulation of disease state.

## Methods

### Clinical Study Design

We conducted an early feasibility trial of adaptive DBS biomarker discovery in adults with severe, treatment-refractory OCD (NCT03457675, NCT04281134 and NCT04806516). The parent clinical trial enrolled ten adults (P1–P10). The trial comprised an initial 6-month open-label DBS programming phase, a 2-month course of exposure and response prevention therapy (months 7–8), a double-blind DBS discontinuation phase (month 9), and open-label follow-up through month 18. The present study represents a secondary analysis of neural and behavioral data acquired during this trial. The first participant was enrolled on September 12, 2018, and the final participant was enrolled on July 25, 2022. A limited subset of data from these patients has been reported previously in studies addressing distinct, unrelated research questions^41,51,52^, and clinical outcomes were previously reported in a separate publication^25^. Information on participant numbers, data collected, and analyses performed across these previous studies is provided in Supplementary Table 1. Each participant gave fully informed consent according to study sponsor guidelines, and all procedures were approved by the local institutional review board at Baylor College of Medicine (H-40255, H-44941, H-49155 to Baylor College of Medicine) and the US Food and Drug Administration (FDA) Center for Devices and Radiological Health (G170268). Participants received a stipend after each study visit via a ClinCard. All participants had a disease duration exceeding five years and had either not responded to or could not tolerate trials of multiple pharmacological treatments (including selective serotonin reuptake inhibitors (SSRIs), clomipramine, and SSRI augmentation with antipsychotics), as well as an expert course of exposure and response prevention (ERP) therapy. We implanted the investigational Activa PC+S system (Medtronic, Inc.) in the first two participants^25^. Of the ten enrolled participants, eight (P3–P10) were eligible for the present longitudinal neurophysiological analyses. We excluded P1 and P2 because the Activa PC+S device used in these participants had limited streaming capabilities in the home environment and therefore did not provide the longitudinal neural recordings required for the present analyses. We implanted the investigational Summit RC + S system (Medtronic, Inc.) in the following eight participants including bilateral DBS leads (Model 3387, Medtronic, Inc.; 1.5-mm contact length and 1.5-mm intercontact spacing) within the ventral capsule (VC), targeted using preoperative MRI^25^. In five of these patients, we additionally placed bilateral four-contact orbitofrontal cortex (OFC) electrocorticography (ECoG) strip electrodes (model 0913025; 4-mm contact diameter and 10-mm intercontact spacing)^25^ to test the hypothesis that cortical signals would improve decodability. In these patients, we implanted two devices per patient, with each system recording from one hemisphere VC and OFC electrodes. We connected the leads to extension cables, tunneled them subcutaneously along the neck, and attached them to the Summit RC + S pulse generator positioned in the upper chest. Throughout the clinical trial, we assessed OCD symptom severity using the Yale-Brown Obsessive Compulsive Scale, Second Edition (YBOCS-II)^5^ during both in-person clinical visits and remote assessments (mean number of YBOCS-II assessments per patient: 22.5±1.5). For the present analysis, we paired YBOCS-II scores with neural recordings acquired during the in-clinic visits. We excluded assessments when neural data was not recorded or contained substantial artifacts (mean number of excluded YBOCS-II assessments per patient: 7.9±5.2). The clinical trial database was locked for analysis on June 10, 2024.

### Pre-specification and statistical power

The parent early-feasibility trials (NCT03457675, NCT04281134 and NCT04806516) prospectively specified the identification of electrophysiological biomarkers of OCD-related distress and DBS-induced hypomania as primary outcomes, with change in OCD symptom severity measured after closed-loop stimulation specified as a secondary clinical outcome. The specific analyses reported in the present study, including comparisons of neural correlates of momentary distress and longitudinal OCD symptom severity, second-order correlation analyses, behavioral feature analyses, and neural representation learning and decoding, were secondary exploratory analyses and were not prespecified trial endpoints. The sample size for these analyses was determined by enrollment in the parent early-feasibility trials and the availability of longitudinal neural recordings rather than by an a priori power calculation. Given the sample size of the parent early-feasibility trials, we report participant-level effect sizes and confidence intervals and, where absence of an association is scientifically interpreted, Bayes factors to quantify evidence for the null hypothesis.

### Electrode localization

We acquired a preoperative clinical computed tomography (CT) scan, T1-weighted (T1w) MRI scan, and a postoperative clinical CT scan to verify electrode placement. We co-registered the postoperative CT to the preoperative T1w MRI and used the aligned images to determine electrode contact locations. We performed automated cortical reconstruction on the preoperative T1w MRI using FreeSurfer v7.1.1^53^, and incorporated the T2-weighted (T2w) MRI to refine pial surface reconstruction. We aligned the postoperative CT to the preoperative T1w MRI using the Linear Image Registration Tool (v6.0) from the FMRIB Software Library^54,55^. We manually identified OFC and VC electrode coordinates from the co-registered CT data in BioImage Suite v3.5b1^56^ and mapped them into native MRI space. Finally, we visualized OFC cortical electrode coordinates using Surfice (https://www.nitrc.org/projects/surfice/) and VC coordinates using Lead-DBS^57^ both in MNI152 standard space.

### Electrophysiological recordings

The Summit RC□+□S system enables simultaneous therapeutic neurostimulation and continuous intracranial local field potential (LFP) recording^58^. We used the Clinician Telemetry Module to establish bidirectional communication between the implanted device and a custom-built application developed with the Research Development Kit, which was connected via Bluetooth to a separate host computer. In both clinical and home settings, we deployed custom clinician- and patient-facing applications on Microsoft Surface Pro and Surface Go tablets. These applications interfaced with the implant and synchronized data to a secure cloud-based storage platform (https://www.box.com/), thereby providing a flexible infrastructure for longitudinal data acquisition and storage. We recorded LFPs at sampling rates of 250, 500, or 1000□Hz during clinic visits and at 250□Hz in the home environment. We configured recordings in a bipolar sensing mode by selecting pairs of adjacent contacts on each lead, referencing the signal from one contact to the other. Supplementary Figure 4 summarizes stimulation contact configurations for each participant. Whenever feasible, we positioned the sensing contact pair to flank the active stimulation contact to minimize stimulation-related artifacts. In addition, we set both stage 1 and stage 2 low-pass filter cutoffs to 100□Hz to further reduce contamination of the recorded signals by stimulation artifacts.

### DBS stimulation parameter programming

At each clinical visit, we obtained two resting-state recordings: one prior to DBS programming and one following parameter adjustments (Supplementary Figure 5). The first resting-state recording reflected the stimulation settings programmed during the previous clinical visit. The second resting-state recording captured the updated stimulation parameters when changes were implemented. We optimized DBS parameters exclusively based on clinical judgment, incorporating standardized clinical assessments (e.g., YBOCS-II), direct interaction with participants, and their subjective reports of mood, anxiety, and alertness during acute programming sessions. To identify optimal stimulation settings, the clinician systematically modified a single parameter at a time, either stimulation amplitude or pulse width. DBS frequency was set to 150.6 Hz for all patients throughout the study.

### Exposure and response prevention therapy recordings

During ERP teletherapy sessions (Figure 1d–f), we instructed participants to initiate a neural recording at the start of each appointment. We saved video recordings locally on the clinician’s computer. Prior to each ERP session, a psychiatrist discussed and defined individualized exposure tasks with the participant. Throughout the session, participants repeatedly rated their distress using the Subjective Units of Distress Scale (SUDS), ranging from 0 to 10.

### Audio and video recordings

During clinical visits, we recorded video and audio using a GoPro Hero 6 at 25 frames per second and audio using a Zoom H4n Pro 4-Track portable recorder at 44.1□kHz. We synchronized the audio and GoPro videos by cross-correlating the video’s onboard audio track with the external audio recording. After alignment, we removed the original GoPro audio and replaced it with the high-quality external audio track. We recorded videos during ERP sessions directly on the Surface Pro tablet and saved the videos with embedded timestamps on each frame. We used the time-of-day information extracted from the video to align the recordings with the corresponding LFP Unix timestamps.

### Electrophysiological signal processing

We used the analysis-rcs-data toolbox to preprocess neural data, account for packet loss, and enable synchronization between LFP and other data streams^59^. For patients with OFC implants, we merged left and right implant recordings based on UNIX timestamps. Within ERP recordings, we aligned SUDS ratings derived from video annotations by converting video time to UNIX timestamps. We annotated and rejected artifacts, including the recently described dual-device beat frequency artifact^51,60^, through manual annotation in 5-second segments. We then computed time-resolved features using the py-neuromodulation feature extraction toolbox^24^. Specifically, we calculated band-power using the scipy Welch function (segment length of 250 samples)^61^. We derived log-transformed band-power across multiple frequency bands: delta (1–4 Hz), theta (4–8 Hz), alpha (8–15 Hz), beta (15–30 Hz), and gamma (30–55 Hz). We used specparam (default parameters, version 1.1.1)^62^ to estimate aperiodic exponent and offset parameters within a fitting range of 10–50 Hz. In addition, we computed bursting features for each of the above-defined frequency bands. We applied the MNE-Python band-pass filter^63^, resampled the signal to 50 Hz, calculated the absolute Hilbert transform, and determined mean burst duration and amplitude using a 75^th^ percentile threshold, as implemented in py-neuromodulation^24,27^. We also extracted temporal waveform-shape features for different filter ranges (1–5 Hz and 1–12 Hz). Waveform-shape features have previously been introduced as complementary LFP characteristics to dissociate physiological and symptomatic states^64–66^. We computed the following temporal waveform shape features in a batch-wise manner for both signal-detected troughs and peaks: prominence, sharpness and interval. Prominence quantifies the mean amplitude difference between a trough and its surrounding peaks (or a peak and its surrounding troughs). Sharpness measures the voltage deflection at each trough or peak relative to voltage values 5 ms before and 5 ms after the detected troughs or peaks, and interval computes the time difference between consecutive troughs or peaks. Because signal polarity can be ambiguous, we computed each waveform-shape feature separately for troughs and peaks and averaged the results. Merk et al. ^24^ described in further detail the computed waveform shape features. Furthermore, we extracted raw LFP voltage amplitudes as well as Hjorth activity, mobility, and complexity parameters, as originally described^67^. We also computed connectivity measures between recording sites. First, we estimated one-second time-resolved aperiodic exponent and offset parameters (see computation method above) and then calculated Pearson correlations between the aperiodic exponent or offset time series of different recording regions. In addition, we computed coherence using scipy^61^ and averaged coherence values within each of the above predefined frequency bands. For ERP recordings, we calculated the mean feature representation surrounding each momentary distress SUDS rating. For in-clinic OCD symptom severity recordings, we computed the average feature representation within each resting-state recording.

### Audio and video signal processing

For both momentary distress and OCD symptom severity, we utilized the same audio and video analysis pipeline. We computed a two-minute average for each SUDS report for momentary distress, and the whole-recording average for clinical interviews captured on the same day as resting-state recordings (Figure 3a). To extract Facial Action Units (FAUs) from video recordings, we first converted the images to grayscale using the OpenCV computer vision library^68^. We then applied the detectMultiScale algorithm (haarcascade_frontalface_default; scale factor = 1.1; min_neighbors = 5) using OpenCV^68^. Next, we extracted FAUs using OpenGraphAU^32^ with a pre-trained ResNet-18 model. Specifically, we extracted the following FAUs: 1, 2, 4, 5, 6, 7, 9, 10, 11, 12, 13, 14, 15, 16, 17, 18, 19, 20, 22, 23, 24, 25, 26, 27, 32, 38, 39, L1, R1, L2, R2, L4, R4, L6, R6, L10, R10, L12, R12, L14, and R14. To compute speech features, we used WhisperX^69^ with the “large-v2” model for transcription. We then applied the default diarization pipeline (version 3.4.2), limiting the output to two speaker identities. We manually corrected all transcription and diarization segments to ensure that only patient speech was included for analysis, and restricted utterances to durations between 2 and 10 seconds. For each patient utterance, we computed a set of speech features. These included embeddings from the speech foundation model wav2vec2^33^ as well as additional signal-processing–based speech features. We hypothesized that these complementary representations would capture speech characteristics relevant for tracking momentary distress and longitudinal symptom severity. From the wav2vec2-large-robust-12-ft-emotion-msp-dim model^31^, we extracted 1,024-dimensional embeddings as well as valence, arousal, and dominance predictions. Using the openSMILE toolbox, we extracted eGeMAPSv02 speech functionals^70^ (n=87) and low-level descriptors^30^ (n=25). A complete list of speech features is available in the openSMILE documentation (https://audeering.github.io/, version 2.6.0).

### Neural decoding pipeline

We separately investigated decoding of momentary distress and OCD symptom severity. Specifically, we examined models trained using neural features alone and evaluated performance improvements when incorporating behavioral variables to learn a joint latent embedding space. We applied the CEBRA framework, which enables both hypothesis-free (neural-only) and hypothesis-guided training approaches^34^. CEBRA is a contrastive learning framework that constructs a low-dimensional latent representation of neural activity by bringing positive pairs closer together and repelling negative pairs according to a supervisory objective.

The framework uses a nonlinear neural network encoder to transform high-dimensional neural features into a latent embedding optimized through contrastive learning. Unlike conventional supervised learning approaches that directly optimize training loss, CEBRA first learns a latent neural representation before fitting a downstream decoder.

In this study, supervisory information consisted of either temporal proximity alone (self-supervised training) or behavioral similarity derived from facial action units and speech features (behavior-guided training). We trained separate CEBRA models for momentary distress and longitudinal OCD symptom severity. Neural features served as inputs to the encoder for all models. During self-supervised training, neural samples that were close in time were encouraged to occupy nearby regions of the latent space. During behavior-guided training, facial and speech features served as auxiliary variables such that neural samples with similar behavioral representations were mapped nearby in the latent space. After training, the encoder weights were frozen, and only neural recordings were used to generate latent embeddings for downstream decoding. Behavioral variables were used exclusively during representation learning and were not provided during model inference or decoding.

We evaluated all decoding analyses using leave-one-sample-out cross-validation. For each fold, a CEBRA encoder was trained exclusively on the training samples, without any temporal overlap to the corresponding test samples, to learn latent neural representations. Separate encoders were trained for momentary distress and longitudinal OCD symptom severity using the hyperparameters described below. After training, the encoder weights were frozen, and latent embeddings were generated for both the training and held-out test samples. Ordinary least squares linear regression models implemented in scikit-learn^71^ were then fit using the training embeddings and corresponding SUDS or YBOCS-II scores and evaluated on the held-out sample. We repeated the retraining process for both the encoder and decoder for each cross-validation procedure. We additionally repeated the training procedure under two conditions: first, by separating behavioral modalities (speech only, FAUs only, or speech and FAUs combined); and second, with different neural feature sets (VC only, OFC only, or both brain regions combined). We evaluated each decoding task using leave-one-sample-out cross-validation. We applied z-score normalization to each sample, fitting the normalization parameters limited to the training data. We reported the Pearson correlation coefficient across all test samples and performed control analyses using shuffled labels.

To predict momentary distress, we used the offset-1 model with the following hyperparameters: learning rate=3e-3, temperature=1.0, conditional=“time_delta”, cosine distance, embedding dimension=50, and hidden units=128. For decoding OCD symptom severity, we used the same offset-1 architecture but adjusted the hyperparameters to account for the smaller sample size: output dimension=8, hidden units=32, conditional=“delta”, and distance=”cosine”. For hypothesis-free training, we did not include behavioral labels and instead used the default “time” conditional.

### Statistical Analysis

We computed all statistical tests using non-parametric Monte Carlo sampled permutation tests with a significance threshold of α=0.05 and *n*=10000 iterations across participants unless otherwise indicated. We used permutation tests to compare paired differences in absolute correlation strength across participants between momentary distress and OCD symptom severity and corresponding brain regions (Figure 2c-e). We reported correlation values and associated p-values using Pearson correlation coefficients computed using scipy (scipy.stats.pearsonr, version 1.16)^61^. We computed p-values for Pearson correlations using a two-sided test against the null hypothesis of zero correlation, based on the exact beta distribution of the sample correlation coefficient, as implemented in scipy. For analyses of correlations between neural features and either momentary distress or OCD symptom severity, we corrected p-values for multiple comparisons across all neural features within each region using the Benjamini–Hochberg method to control the false discovery rate (statsmodels.stats.multitest.multipletests version 0.14.5)^72^. To assess whether differences between neural OCD symptom severity and momentary distress correlations were consistent across participants, we additionally calculated within-participant differences. We estimated group-level 95% confidence intervals by bootstrap resampling with replacement. To quantify evidence for the null hypothesis, we additionally computed Bayes factors for second-order Pearson correlations using Bayesian one-sample t-tests with a default Jeffreys–Zellner–Siow Cauchy prior on the standardized effect size with scale r=0.707 (implementation pingouin.bayesfactor_ttest in pingouin)^73^. We reported Bayes factors as BF_01_, representing the likelihood of the observed data under the null relative to the alternative hypothesis. For assessing the significance of the number of neural features across TEED stimulation parameter changes, we used Bernoulli tests to compare the number of significant neural feature changes across conditions: ΔTEED=0 within clinical visits, ΔTEED=0 across clinical visits and ΔTEED≠0 within clinical visits. We performed all decoding analyses using within-subject leave-one-sample-out cross-validation analysis. All CEBRA encoders and least-squares regression decoders were thus retrained for each fold to predict left-out samples. We compared decoding performance against shuffled label performances, using a defined random seed. We assessed significance of decoding performances using “time” against behavior-guided learning using relative permutation tests. In a similar analysis we assessed statistical comparisons within regions using relative permutation tests using behavior-guided (FAUs + speech) against “time” self-supervised distributions. We applied participant-level bootstrap and sign-flip permutation procedure to compare neural-only CEBRA decoding performance with decoding using FAU and speech features as behavioral auxiliary variables. All data are reported as mean ± standard deviation unless otherwise noted. All analyses were conducted in Python 3.12 and MATLAB R2024b. Software versions are accessible in the pyproject.toml version control files within the publicly available code repository.

## Data Availability

All data produced in the present study are available upon reasonable request to the authors.

## Code and data availability

The data that support the findings of this study are available from the corresponding author upon reasonable request. All code is publicly available on GitHub: https://github.com/ProvenzaLab/multiscale-psychiatric-neural-state-representations

## Supplementary Tables

**Supplementary Table 1:** Participant-level overlap with previous publications. Participants were enrolled in the parent early-feasibility DBS study. Participant identifiers are consistent across publications. Previous publications used subsets of the cohort and/or data to address distinct research questions.

| Participant ID(s) | Previous publication | Previously reported data/analysis |
| --- | --- | --- |
| P1,P2,P3,P4,P5 | Provenza et al., *Nat Med* (2021) | Initial five-participant cohort. Analysis of correlations between canonical frequency-band activity in the ventral capsule (VC) during ERP and SUDS. Technical demonstration of simultaneous neural and behavioral recordings during ERP. |
| P6,P7,P8 | Alarie et al., *Front Hum Neurosci* (2022) | Characterization and mitigation of stimulation-related artifacts in Summit RC+S recordings from three OCD participants. |
| P10 | Provenza et al., *J Neurosci* (2025) | Single-participant analysis of vIPFC high-beta activity, DBS amplitude and approach behavior/speech. |
| P10 | Herron et al., *Nat Biomed Eng* (2025) | Perspective on cortical sensing for adaptive DBS; includes illustrative chronic OCD recordings of a single participant. |
| All participants (P1-P10) | Sheth et al., preprint (2026) | Clinical outcomes paper: The study evaluated clinical efficacy, safety, neuropsychological outcomes, and the feasibility of chronic intracranial sensing across the 18-month early-feasibility trial. |

**Supplementary Table 2:** Mean participant-level neural Pearson correlations with OCD symptom severity and momentary distress and their respective differences.

| Participant ID | OCD symptom severity neural correlations | Momentary distress neural correlations | Neural correlation difference |
| --- | --- | --- | --- |
| 3 | 0.488 | 0.212 | 0.276 |
| 4 | 0.444 | 0.222 | 0.222 |
| 5 | 0.425 | 0.105 | 0.321 |
| 7 | 0.582 | 0.202 | 0.379 |
| 8 | 0.516 | 0.306 | 0.21 |
| 9 | 0.233 | 0.178 | 0.056 |
| 10 | 0.375 | 0.486 | -0.111 |

**Supplementary Table 3:** Mean participant-level neural Pearson correlations with OCD symptom severity and momentary distress separated for OFC and VC regions and differences across regions.

| Measure | Participant ID | OFC neural correlation | VC neural correlation | Neural correlation difference |
| --- | --- | --- | --- | --- |
| Momentary distress | 7 | 0.251 | 0.143 | 0.107 |
|  | 8 | 0.301 | 0.295 | 0.006 |
|  | 9 | 0.195 | 0.16 | 0.035 |
|  | 10 | 0.54 | 0.287 | 0.252 |
| OCD symptom severity | 6 | 0.677 | 0.439 | 0.238 |
|  | 7 | 0.593 | 0.514 | 0.079 |
|  | 8 | 0.587 | 0.354 | 0.232 |
|  | 9 | 0.249 | 0.205 | 0.043 |
|  | 10 | 0.418 | 0.297 | 0.12 |

**Supplementary Table 4:**
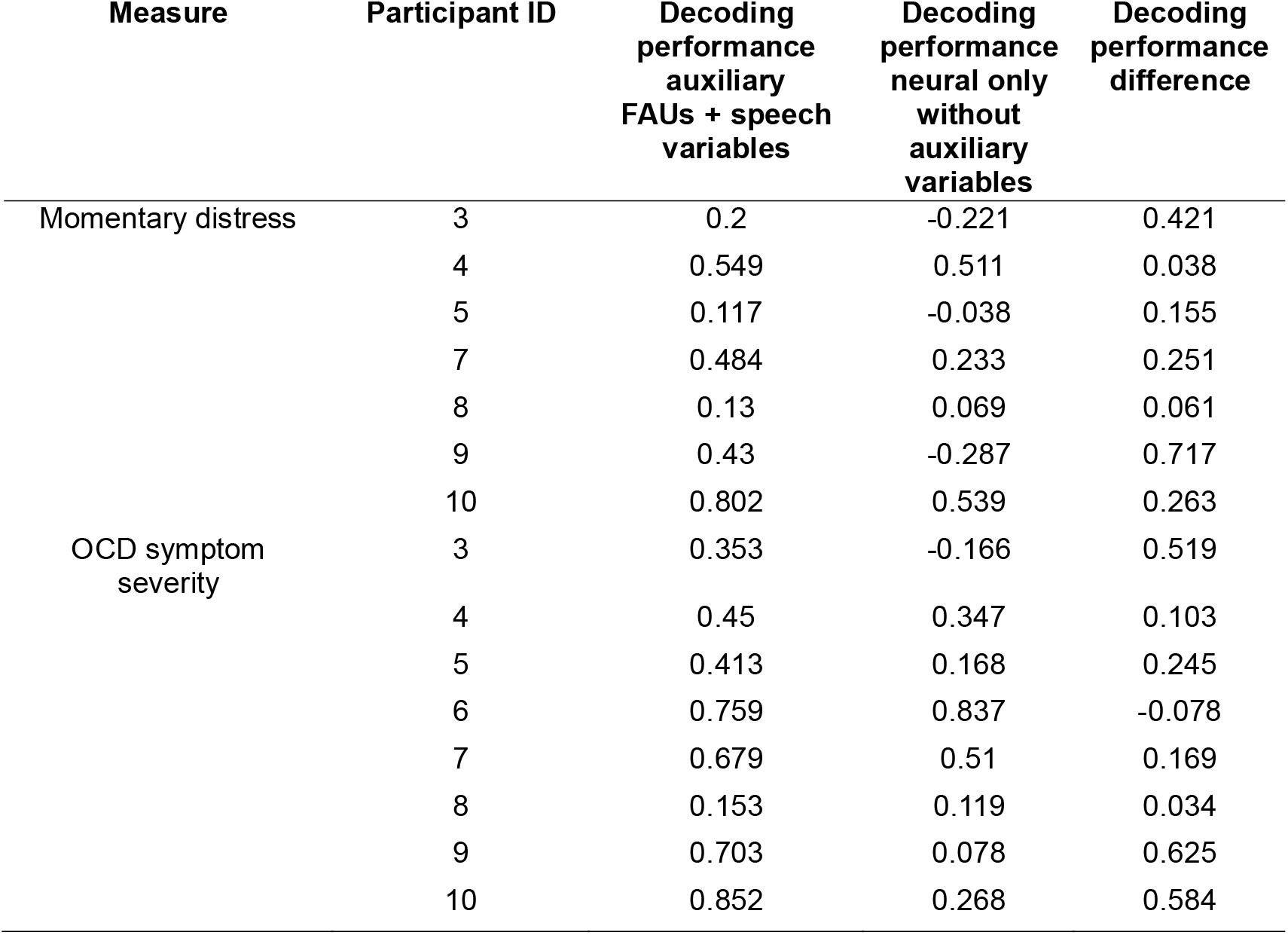
Mean participant-level decoding performances separated for FAUs + speech auxiliary variables vs neural only decoding.

## Supplementary Figures

**Supplementary Figure 1.**
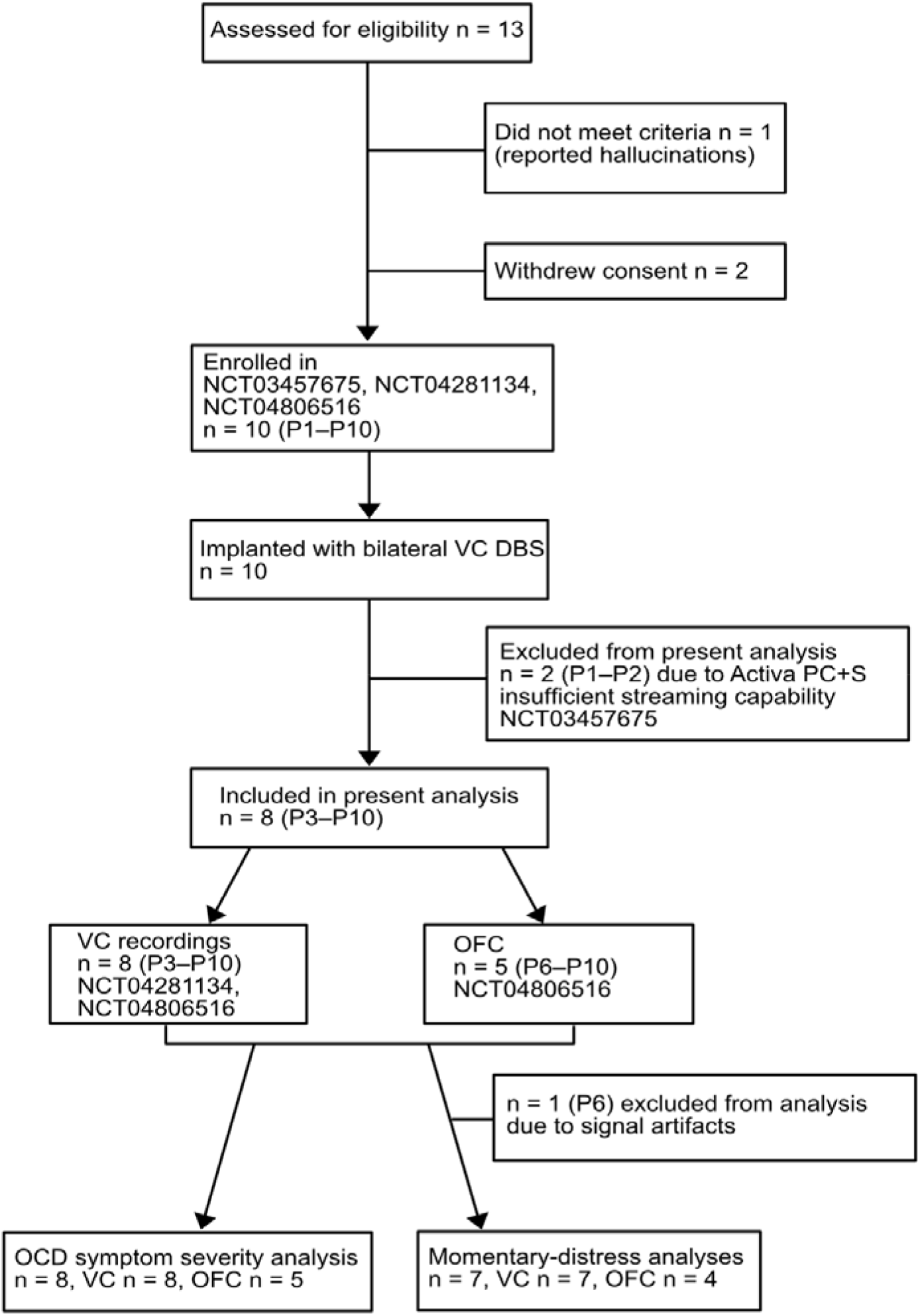
Participant flow diagram. Participant inclusion summary in the present neural and behavioral analyses. Ten participants were enrolled in the clinical trial, of whom eight contributed data to the analyses reported here. Primary clinical outcomes of the trial have been reported previously^25^.

**Supplementary Figure 2.**
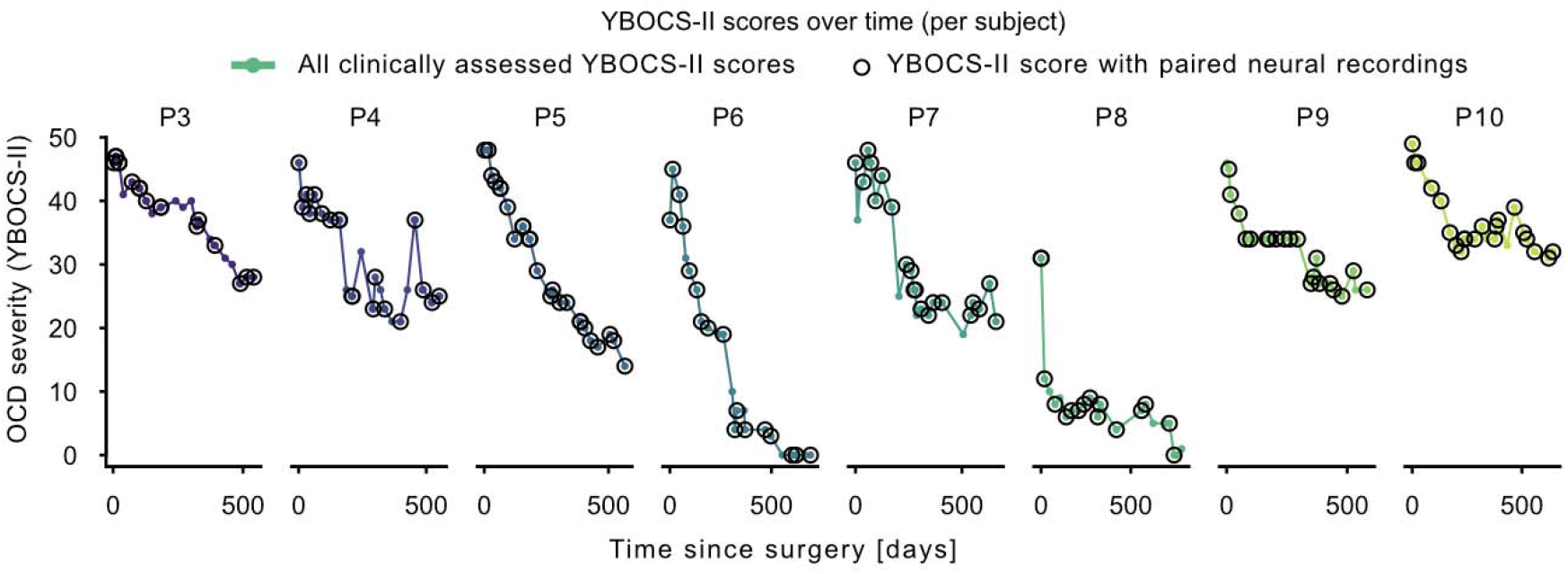
OCD symptom severity fluctuations across patients since surgery. All clinically recorded YBOCS-II ratings are displayed, with timepoints included in this analysis indicated by circles. Not all OCD symptom severity assessments were paired with neural recordings, as some were conducted remotely without neural recordings or were excluded due to recording artifacts.

**Supplementary Figure 3.**
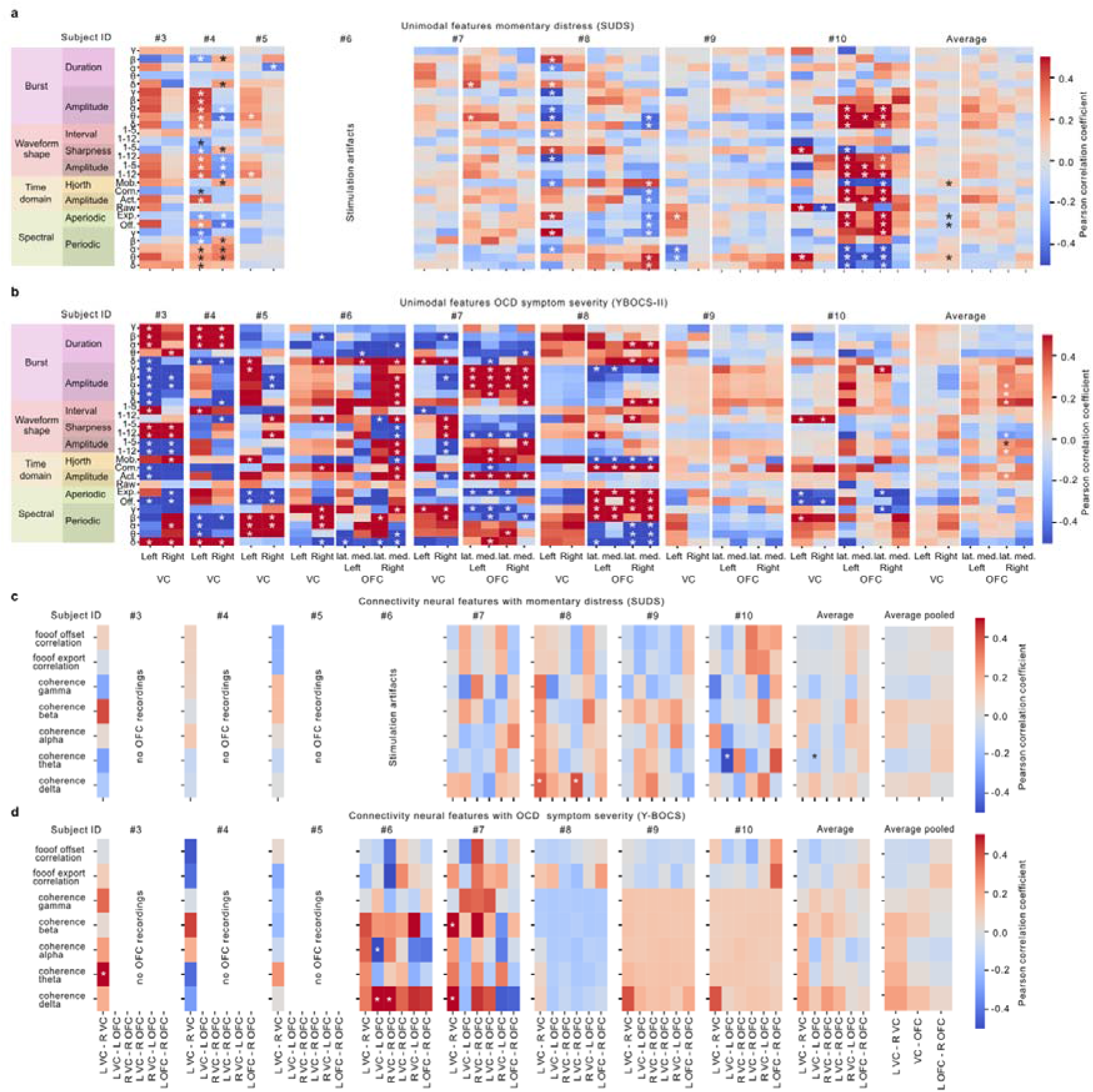
Patient-specific neural feature correlation maps. (a,b) Univariate neural feature correlations with momentary distress (a) and longitudinal OCD symptom severity (b). (c,d) Connectivity feature correlations with momentary distress (c) and longitudinal OCD symptom severity (d). Significant correlations after multiple-comparison correction are indicated by (*).

**Supplementary Figure 4.**
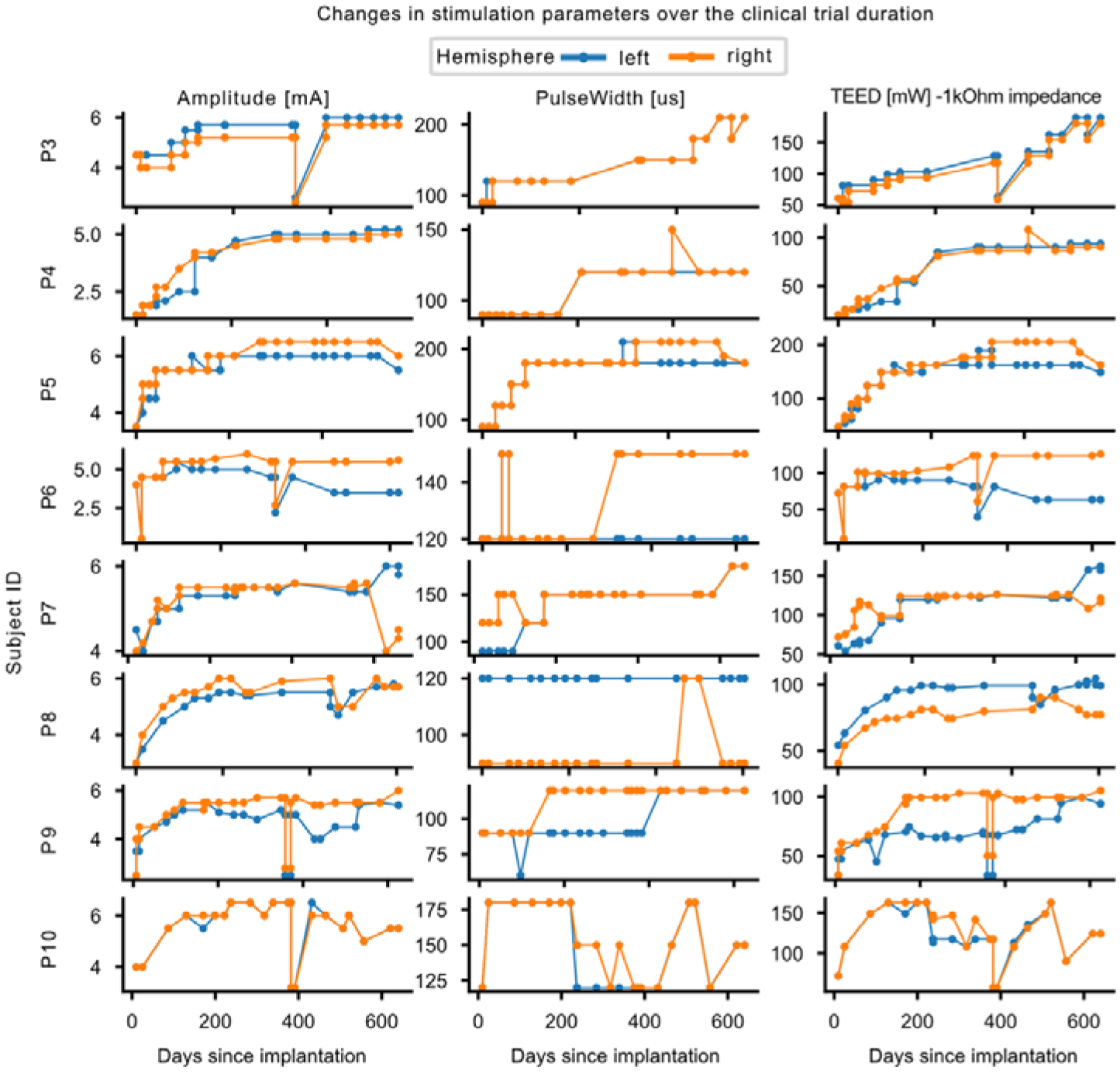
Patient-specific DBS parameter changes across the clinical trial. Stimulation amplitude, pulse width, and total electrical energy delivered (TEED; assuming a constant 1-kΩ impedance) are shown separately for the left and right hemispheres across the study duration for each patient.

**Supplementary Figure 5.**
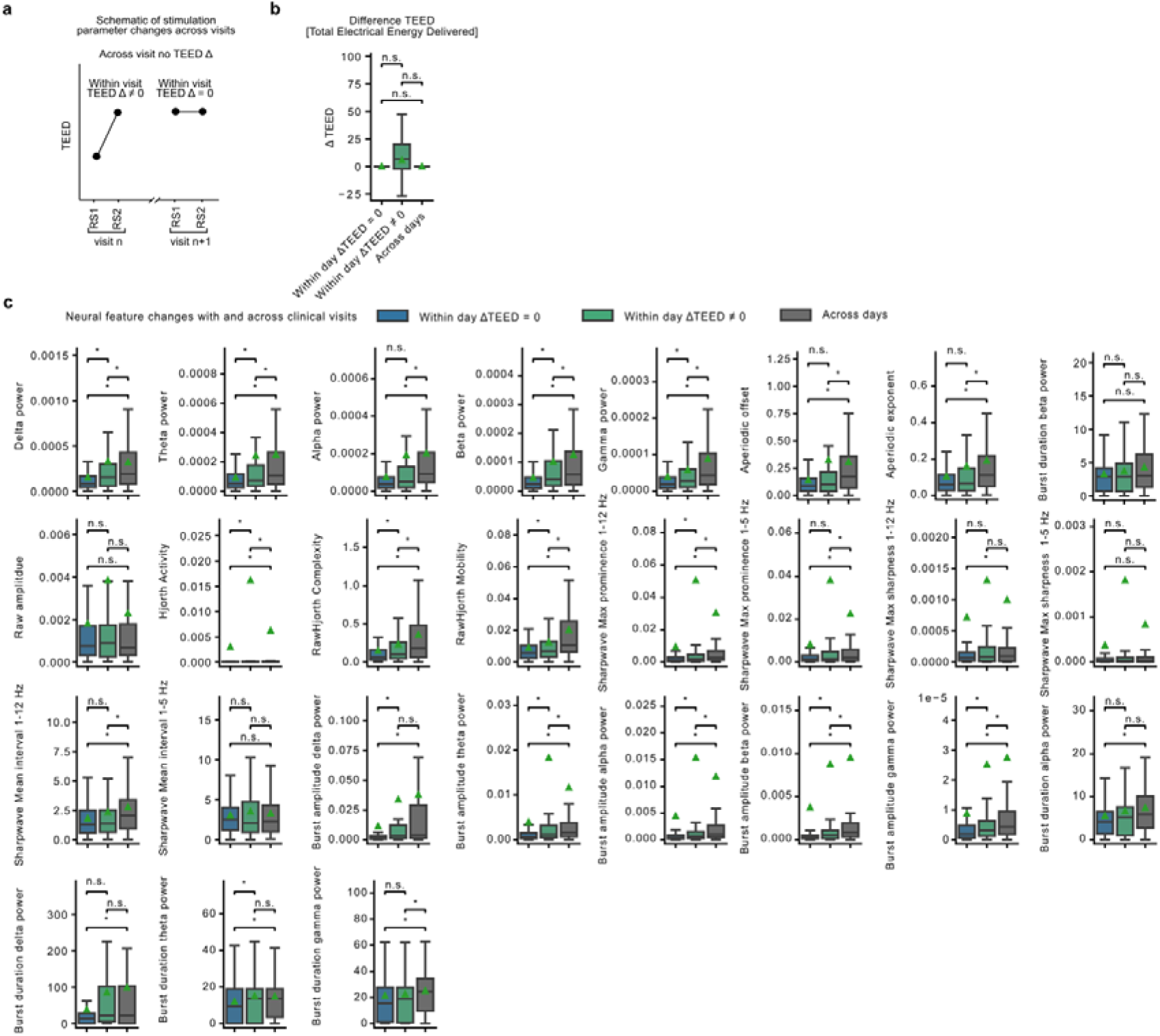
Neural feature changes within and across clinical visits. (a) At each clinic visit, resting-state (RS) recordings were acquired immediately before and after DBS programming, creating a within-visit control that isolated the acute effects of stimulation parameter adjustments from longitudinal changes in clinical state. Changes in neural features observed within visits (*RS1_t_* and *RS2_t_*) were then compared with those observed across consecutive visits (*RS2_t-1_* and *RS1_t_*). (b) Because stimulation parameters were adjusted only during clinical visits, recordings acquired across consecutive visits were obtained under identical stimulation settings, with total electrical energy delivered (TEED) remaining unchanged (ΔTEED=0), whereas within-visit comparisons captured neural activity both before and after DBS programming changes. (c) Distributions of changes in individual neural features shown for three comparisons: within-visit recordings before and after DBS programming when stimulation parameters changed (blue), within-visit recordings without stimulation parameter adjustments (green), and across consecutive clinic visits acquired under identical stimulation settings (gray). Although DBS programming produced measurable changes in neural features, longitudinal changes observed across visits were significantly greater than within-visit changes regardless of whether TEED changed (ΔTEED=0: Bernoulli test number of significant changes across features *p*=1.7·10^−26^; ΔTEED≠0: *p*=1.5·10^−17^).

**Supplementary Figure 6.**
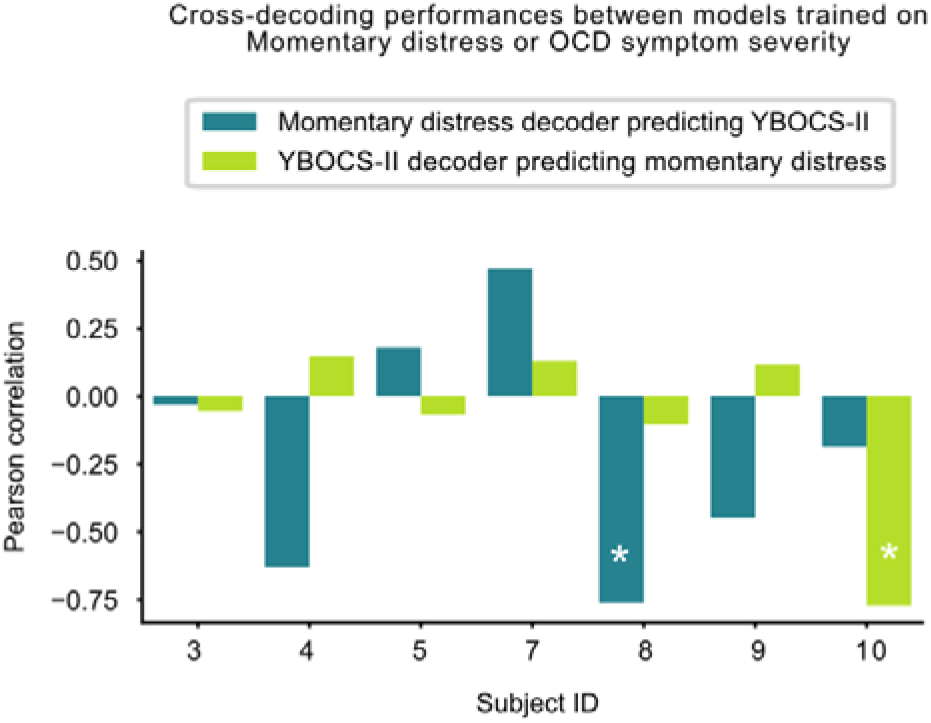
Cross-decoding analysis between momentary distress and longitudinal OCD symptom severity. Participant-specific cross-decoding performances for models trained to predict momentary distress and tested on longitudinal OCD symptom severity (blue), and trained on longitudinal OCD symptom severity and tested on momentary distress (green).

## Acknowledgements

This work relied heavily on the community expertise and resources made available by the Open Mind Consortium (https://openmind-consortium.github.io/). Summit RC + S systems were donated by Medtronic, Inc. as part of the BRAIN Initiative Public-Private Partnership Program. This research was supported by the National Institutes of Health (NIH) National Institute of Neurological Disorders and Stroke BRAIN Initiative via contract UH3NS100549 (to S.A.S., W.K.G., N.R.P., and E.A.S.), the National Institutes of Health (NIH) National Institute of Mental Health (NIMH) R01MH139889 (N.R.P.) and NIH BRAIN Initiative via UH3NS136631 (W.K.G., N.R.P., S.A.S.), the McNair Foundation (S.A.S., N.R.P.), the Gordon and Mary Cain Pediatric Neurology Research Foundation (S.A.S.) and Brain and Behavior Research Foundation Young Investigator Award (N.R.P).

## Contributions

T.M., N.R.P. and S.A.S. conceived the study. T.M. conceptualized data analysis procedures, performed data analysis, interpreted data and prepared figures and results, with support from N.R.P., S.A.S., G.B., R.H., B.R., H.Y., J.A., M.O., T.H., V.B., S.S., G.R., S.V.R., and T.F. A.D.W. performed data collection during ERP therapy. M.A.-O., W.K.G., N.R.P., and S.V.R. performed data collection in the clinic. T.M. wrote the first draft of the manuscript, and all authors contributed to the writing and revision of the manuscript. W.K.G., S.A.S., and E.A.S. performed the clinical care aspects of the study. S.A.S. performed the study surgical procedures. N.R.P. oversaw data collection, analysis and manuscript completion.

